# A Quantitative Characterization of the Spatial Distribution of Brain Metastases from Breast Cancer and Respective Molecular Subtypes

**DOI:** 10.1101/2022.07.05.22277116

**Authors:** Saeedeh Mahmoodifar, Dhiraj J. Pangal, Tyler Cardinal, David Craig, Thomas Simon, Ben Yi Tew, Wensha Yang, Eric Chang, Min Yu, Josh Neman, Jeremy Mason, Arthur Toga, Bodour Salhia, Gabriel Zada, Paul K. Newton

## Abstract

1.

Brain metastases (BM) remain a significant cause of morbidity and mortality in breast cancer (BC) patients. Specific factors promoting the process of BM and predilection for selected neuro-anatomical regions remain unknown, yet may have major implications for prevention or treatment. Anatomical spatial distributions of BM from BC suggest a predominance of metastases in the hindbrain and cerebellum. Systematic approaches to quantifying BM location or location-based analyses based on molecular subtypes, however, remain largely unavailable. We analyzed stereotactic Cartesian coordinates derived from 134 patients undergoing gamma-knife radiosurgery (GKRS) for treatment of 407 breast cancer BMs to quantitatively study BM spatial distribution along principal component axes and by intrinsic molecular subtype (ER,PR,Herceptin). We corroborated that BC BMs show a consistent propensity to arise posteriorly and caudally, and that Her2+ tumors are relatively more likely to arise medially rather than laterally. To compare the distributions among varying BC molecular subtypes, we used the notion of mutual information, which revealed that the ER-PR-Her2+ and ER-PR-Her2-subtypes showed the smallest amount of mutual information and were most molecularly distinct. Using kernel density estimators, we found a propensity for triple negative BC to arise in more superiorly or cranially situated BMs. BM location maps according to vascular and anatomical distributions using cartesian coordinates to aid in systematic classification of tumor locations were additionally developed. Further characterization of these patterns may have major impacts on treatment or management of cancer patients.

**Significance:** The quantitative spatial distribution of breast cancer metastases to the brain, and the effects of breast cancer molecular subtype on distribution frequencies remain poorly understood. We present a novel and shareable workflow for characterizing and comparing spatial distributions of BM which may aid in identifying therapeutic or diagnostic targets and interactions with the tumor microenvironment.

## 2. Introduction

In patients with breast cancer (BC), brain metastases (BM) are a significant source of morbidity and mortality, and average interval between diagnosis of BM and death remains under two years16. Despite significant advances in systemic treatment of primary breast cancer, treatment for BM remains mostly confined to surgical resection, stereotactic radiosurgery, and less commonly whole brain radiation therapy.

BM from BC have been reported to show preferential spatial metastatic patterns within the brain, with a predominance of lesions arising in the posterior circulation and cerebellum [1–3].While the spatial distributions for BM have been described in a qualitative fashion (e.g. located in cerebellum), there have been minimal efforts to systematically and quantitively analyze spatial distributions of BM. In addition, the influence of molecular subtype in topographic BM distribution remains largely unknown.

There is relevant clinical and potential therapeutic motivation for understanding the spatial distribution of BM, specifically according to cancer origin and molecular subtype. There has been growing interest in the relationship between the tumor microenvironment (TME), surrounding both tumor and normal brain parenchyma, and the development of BM, which is referred to as the ‘seed-and-soil’ hypothesis [4–6]. Recent studies have characterized a need for priming a metastatic niche prior to BM colonization and tumorigenesis [7–9]. A more thorough understanding of the patterns of spatial distribution of BM and the influence of TME on tumorigenesis may provide potential targets for diagnosis or treatment of BM.

Gamma Knife Radiosurgery (GKRS) is a highly targeted form of stereotactic radiosurgery and is a first line therapy for many BM, particularly those in which surgical resection is unfavorable [10,11]. The use of stereotactic frames and precise, predetermined locations in three-dimensional space allow for Cartesian coordinates of tumors to be recorded and studied (Figure 1) using spatial modeling techniques. We describe a novel computational approach for characterizing and comparing the spatial distribution of BM arising from BC, using objective tumor location data from patients undergoing GKRS. Tumor locations were analyzed using kernel density plots and principal components analyses (data-based coordinates), and further characterized and compared according to BC molecular subtype. We compared two distributions using the metric of mutual information which is a (nonlinear) measure of the mutual dependence between two random variables [12]. A standard interpretation of mutual information is that it quantifies the amount of information obtained about one random variable by observing the other, thus low values indicate that the distributions are more distinct (independent) than distributions with higher values. While this study introduces new tools for quantifying spatial distributions of BM using large comprehensive data sets collected over a twenty-year period, it also paves the way for further analyses with larger, prospective multi-center studies across a variety of cancers and molecular subtypes to further elucidate natural distribution patterns of BM and their importance for improving cancer treatment.

**Fig. 1.**
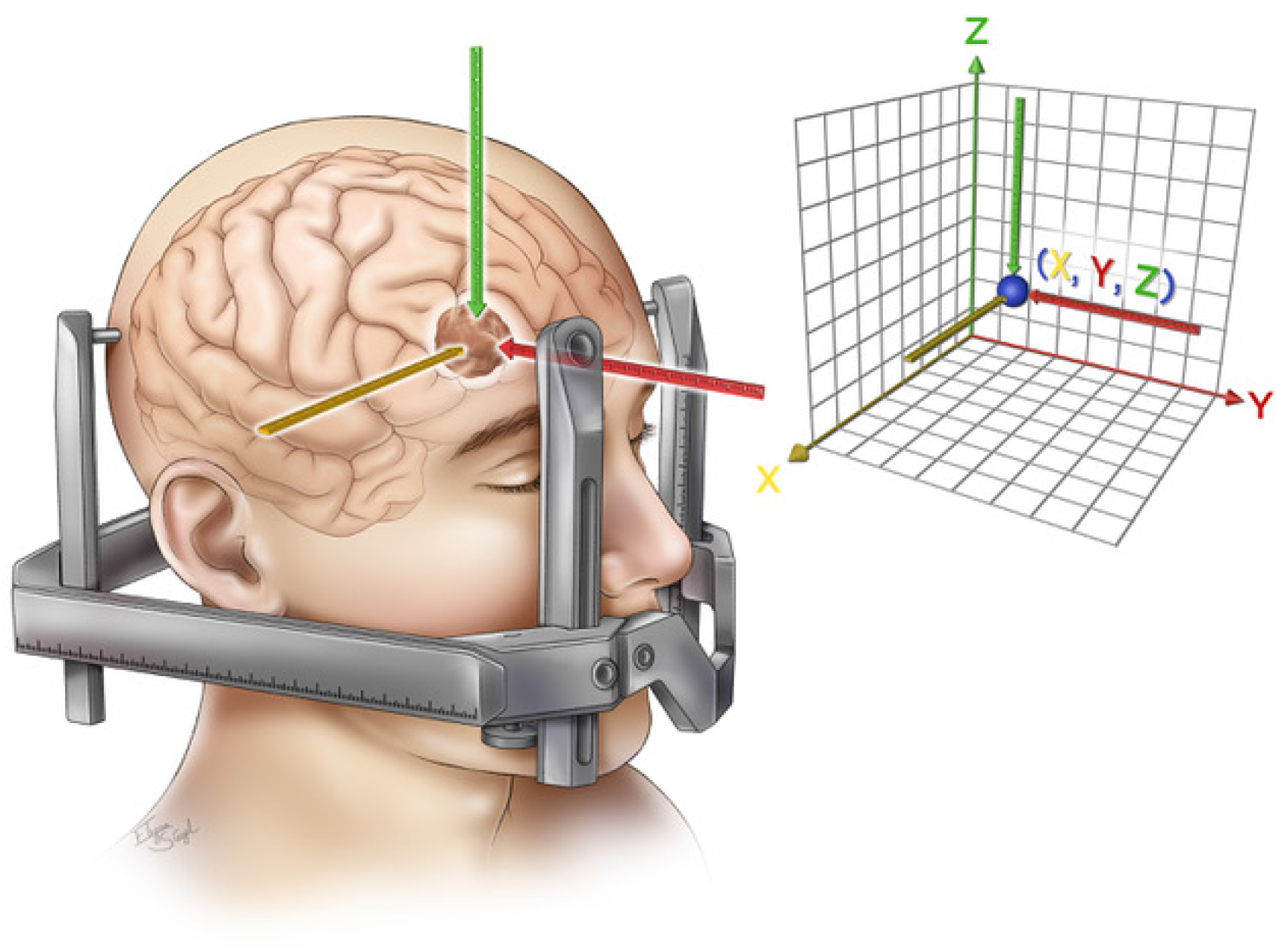
Schematic illustration of Gamma Knife radiosurgery (GKRS) stereotactic headset, intracranial metastasis (brown), and targeted radiation location in X (yellow), Y (Red) and Z (Green) planes. These coordinates are subsequently mapped to a traditional three dimensional cartesian plane (right), and repeated for all brain metastases for all patients undergoing GKRS at our institution.

## 3. Methods

### 3.1 Radiosurgery Setup and Patient Selection

Gamma Knife radiosurgery (GKRS) is a commonly used frontline treatment modality in which a stereotactic frame (Leksell coordinate frame, see Figure 1) is used in conjunction with cobalt radiation sources to deliver precise doses of radiotherapy to highly accurate locations in three-dimensional space corresponding to contoured BM on MRI (Figure 2). Predetermined target coordinates are utilized, and patients are fixed to the stereotactic Leksell coordinate frame as depicted in Figure 1. As a result, Cartesian coordinates (X,Y,Z) in 3D space of each BM central location are obtained and recorded.

**Fig. 2.**
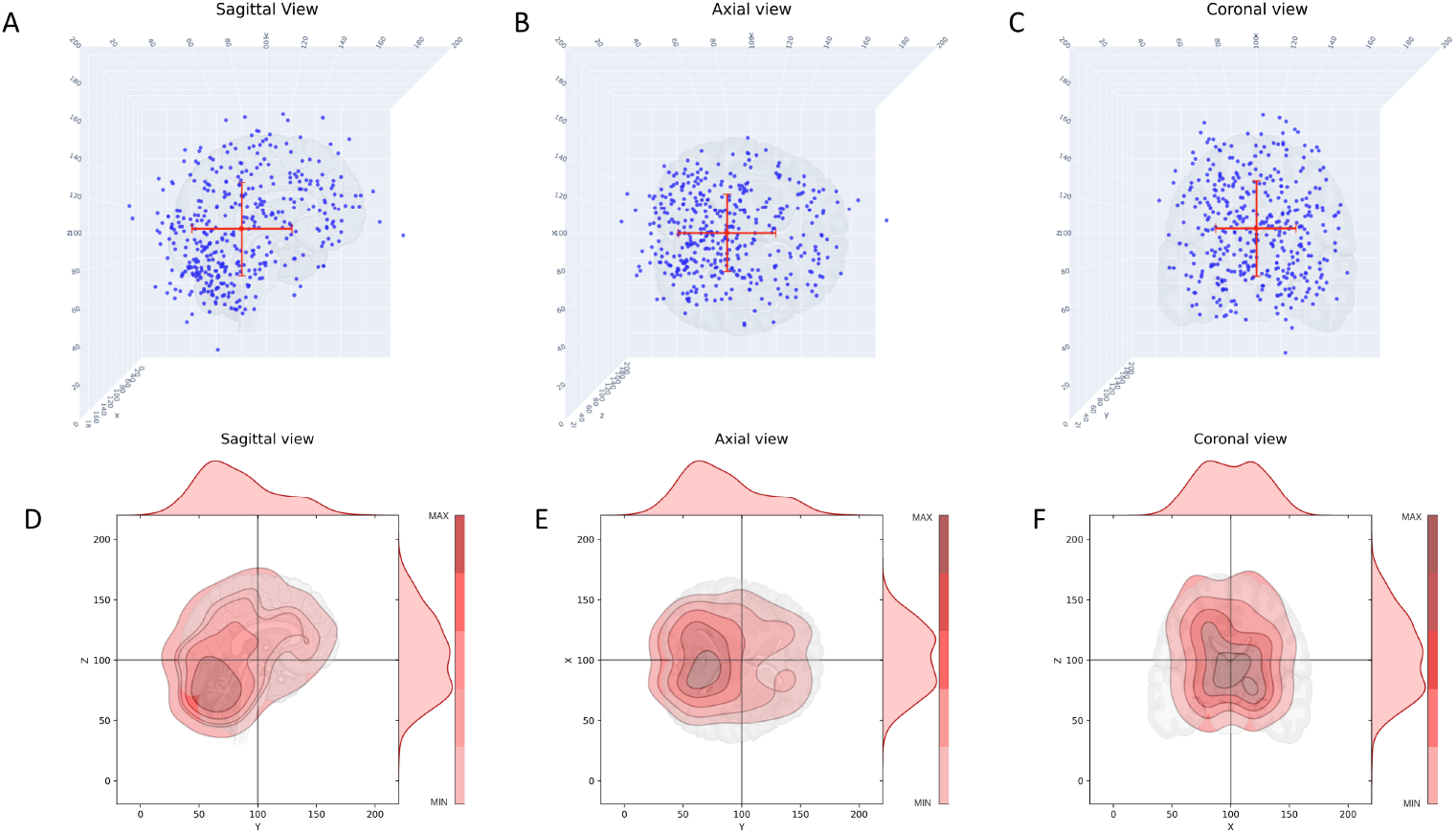
Scatter and kernel density plots showing the spatial distribution of metastatic brain tumors for all breast cancer patients in sagittal, axial, and coronal views. A) Scatter plot, sagittal view, red dot indicates the mean; B) Scatter plot, axial view, red dot indicates the mean; C) Scatter plot, coronal view, red dot indicates the mean; D) Kernel density plot, sagittal view. Color shading indicates density, closed dark regions indicate highest density of metastatic tumors. Distributions on top and right are probability distribution functions (pdf’s) describing the distribution of tumors. E) Same as (D), axial view; F) Same as (D), coronal view.

All patients undergoing GKRS at The Keck Hospital of the University of Southern California (USC) between the years 1995-2015 for the treatment of BM were reviewed and analyzed following approval from the local USC IRB. Those with primary BC were identified, and retrospective chart review was conducted to determine molecular subtype (ER, PR and Her2/Neu [HER2]). Samples were divided into 6 major subtypes based on HER2, ER and PR receptor status. Subtype information was available in 134 patients comprising a total 407 intracranial metastases. Clinical data gathered included: sex, age at diagnosis of primary cancer, age at diagnosis of BM, ER status, PR status and Her2/Neu status. To avoid potential confounders with prior radiation therapy, only patients with their first radiation treatment were included and those with prior radiation or radiosurgery were excluded. Multiple metastases from individual patients (at one treatment) were included. See data summary in Table **??**.

GKRS planning and treatment were performed by a multidisciplinary team including a neurosurgeon, radiation oncologist, and medical physicist. Tumor locations were recorded as (X,Y,Z) values on a Cartesian plane, corresponding to the Leksell coordinate frame axes and recorded using GammaPlan ™ software (Elekta corporation). In addition, specific clinical locations (e.g. Left frontal lobe) as well as tumor volume, number of treatments, vascular distribution, and radiation dose were recorded.

### 3.2 Principal component analysis (PCA) and mutual information (MI)

The principal component (PC) coordinates are a data-based orthogonal coordinate system designed to bring out the directions of maximal spread of the data and used in many settings in which patterns are sought from large data sets [13]. The PC coordinates are linear combinations of the three (X,Y,Z) physical coordinates, with mean at the origin, mutually orthogonal (so they span the same space as X-Y-Z), and such that PC1 lies in the direction of maximal spread, PC2 is orthogonal to PC1 and is in the next most likely direction of spread, while the PC3 direction is orthogonal to both, with the least direction of spread. Since the method of calculating the PC coordinates is standard, we refer the interested readers to Kirby [13] for theoretical details. We use scikit-learn Python package [14] for our data analysis. To compare two distributions associated with different molecular subtypes, we use the notion of mutual information (MI) [12] (relative entropy) which quantifies nonlinear mutual dependence between two random variables. If the MI is zero between two random variables, they are deemed to be completely independent and unrelated, which implies that using observations drawn from one has no value in predicting sequences generated by the other. The formula we use to estimate MI is [15]:

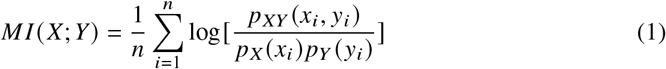

where *P*_*XY*_ (*x*_*i*_, *y*_*i*_) is the estimated joint PDF, and *P*_*X*_ (*x*_*i*_) and *P*_*Y*_ (*y*_*i*_) are the estimated marginal PDF’s at (*x*_*i*_, *y*_*i*_). The larger the MI value, the more the distributions are correlated, i.e. one distribution carries a high amount of information about the other. A very useful discussion and application of MI can be found in reference [16].

### 3.3 Kernel density estimators and bootstrap method

Kernel density estimators offer a useful tool to convert a discrete multivariate data set into smoothed, multivariate distributions to extract information and patterns associated with the probability distribution function associated with data [17]. Color gradient bars and contours are then used to identify ‘hot spot’ regions of highest density (probability), and regions of lowest density (probability). In principle, the computed MI does not depend on the size of the data sets being compared, although well known issues can arise from smaller data sets [15, 16]. For these reasons, to overcome the issue associated with small and unequal sizes of data sets for different molecular subtypes, we use a bootstrap (resampling) method [18], starting from the smoothed multivariate distributions obtained for each subtype (from the original data sets) to generate sample data of 1000 points and then calculate the MI values (see Table S2) for those points between each pair of subtypes. We carry out this re-sampling step and MI calculation step 1000 times, and obtain sample means and standard deviations for the MI for each pair using the enlarged data sets generated from sampling from the distributions generated from the original data sets.

## 4. Results

The data set is compiled in Table **??** which shows the number of BM for each of the molecular subgroups, as well as details associated with Figures 2-8, S1, S2. Figure 2 shows the entire data set of brain metastases (Figure 2 A,B,C) for our cohort of breast cancer patients, in the sagittal, axial, and coronal planes. These same views are shown in Figure 2 D,E,F as kernel density plots depicting the density distributions associated with the data. The darkest enclosed regions of the kernel density plots nicely depict the highest density regions (‘hotspots’), which generally cluster towards the midline (coronal, axial view), posteriorly and caudally (sagittal). Figure S9 shows the same data broken down according to the molecular subtype (along each column): ER-PR-Her2+; ER+PR+Her2-; ER-PR-Her2-(TNBC); ER+PR+Her2+ (TPBC). The red dot marks the mean position. The corresponding kernel density plots for the molecular subgroups are shown in Figure 3. The sagittal view across all subtypes (Figure 3, Row 1) demonstrates clear maximal clustering in the posterior, caudal region of brain; however TNBC appears to visually cluster superiorly/cranially compared to the other breast cancer subtypes. We next focused on elucidating differences in topographic patterns associated with the molecular subgroups by using the principal component axis coordinates [13]. The principal component coordinates are a rotated orthogonal coordinate system centered at the mean of the data that are optimally designed to highlight the largest spread direction (PC1). In Figure 4 we show the relationship between the principal component coordinates (PC1-PC2-PC3) and the physical cartesian coordinates (X-Y-Z). Figure 4 A shows PC1-PC2-PC3 in the X-Y-Z space, while Figures 4 B,C,D shows each of the two-dimensional projections. From Figure 4 B we can see that PC1 lies predominantly in the anterior-posterior (Y), although with other components as well (Figure 4 C,D). The precise linear relationship between the two coordinate systems is given by:

**Fig. 3.**
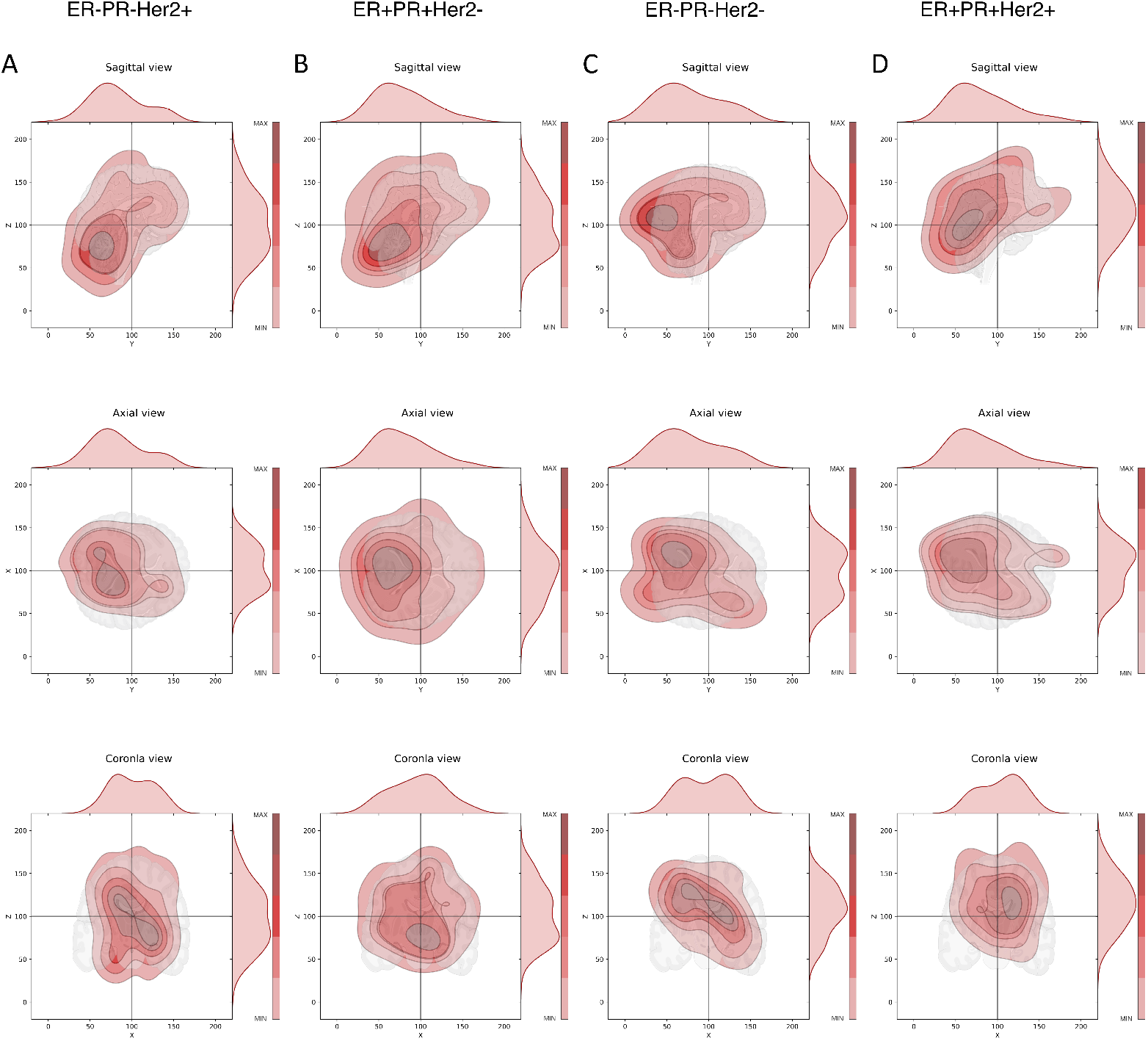
Kernel density plots of metastatic tumor distributions according to genetic subgroups, sagittal, axial, and coronal views. Color shading indicates density, closed dark regions indicate highest density of metastatic tumors. Distributions on top and right are probability distribution functions (pdf’s) describing the distribution of tumors. Column showing ER-/PR-/Her2+ subgroup, three views; B) Column showing ER+/PR+/Her2-subgroup, three views; C) Column showing TNBC subgroup, three views; D) Column showing TPBC subgroup, three views.

**Fig. 4.**
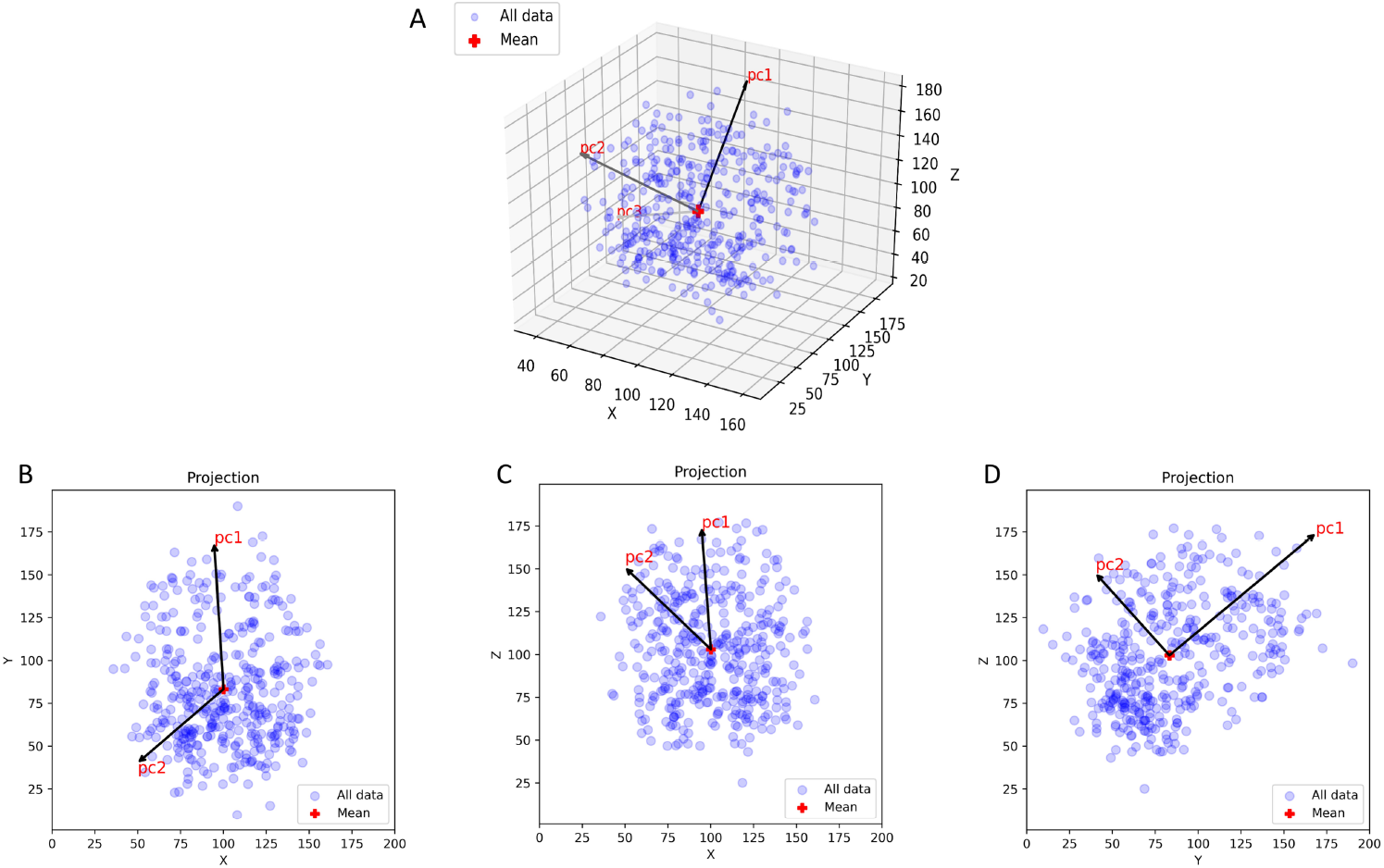
Scatter plot of all breast cancer metastatic brain tumors in X-Y-Z coordinates showing the Principal component axes PC1-PC2-PC3. A) 3D data representation in (X,Y,Z) space showing the orientation of (PC1,PC2,PC3). B) 2D projection onto (X,Y) plane; C) 2D projection onto (X,Z) plane; D) 2D projection onto (Y,Z) plane.

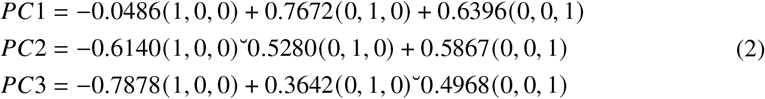

In Figure S10 we compare the spatial distributions in the original X-Y-Z coordinates and the principal component axes (PC1-PC2-PC3) from the full data set for the six molecular subtypes: Her2+, ER+, PR+, PR-, Her2-, ER-separately. In each plot, the yellow horizontal bar marks the mean, while the white dot marks the median. The colors mark the molecular subtype, as shown in Figure S10 A which most clearly shows the divergence along the PC1 axis which is the direction of maximal spread. To understand the advantages of using the principal component coordinates over the cartesian coordinates, in Figure S10 A it is clear that the median lies below the mean (i.e. is shifted back with respect to the mean), with the three negative subtypes shifted further back than the three positive ones. Comparing this with Figure S10 E (spread along Y-axis), the pattern is not nearly as clear. For each pair of violin plots (distributions), we calculate the mutual information score (MI) along with standard deviations using the bootstrap method described earlier. Lower MI score indicates less mutual dependence between the compared distributions, higher MI score indicates more mutual dependence. Figure 5 A-F shows the same as Figure S10, but using the molecular subgroupings: TPBC; ER+PR+Her2-;ER-PR-Her2+; TNBC. The divergence between the mean and the median is largest in the triple negative grouping, shown most clearly in Figure 5 A along the PC1 axis. An ordered listing of all of the MI scores for each pair of molecular subtypes is shown in Table S2 and presented visually for the individual subtypes in Figure 8 as a heat map. The ordering in Table S2 goes from smallest to largest along the PC1 axis (first column), with all other axes also shown. In Table S2 and Figure 5 A we draw attention to the fact that the pair with the smallest MI value (8.966 +/-3.394) is between ER-PR-Her2+ and ER-PR-Her2-, i.e. those two groupings are the most molecularly distinct. The two groups with the largest MI value (14.808 +/-3.589) is between ER+PR+Her2+ and ER+PR+Her2-, i.e. those two groupings are the most molecularly similar (more important than the nominal values of these MI scores are the differences between them).

**Fig. 5.**
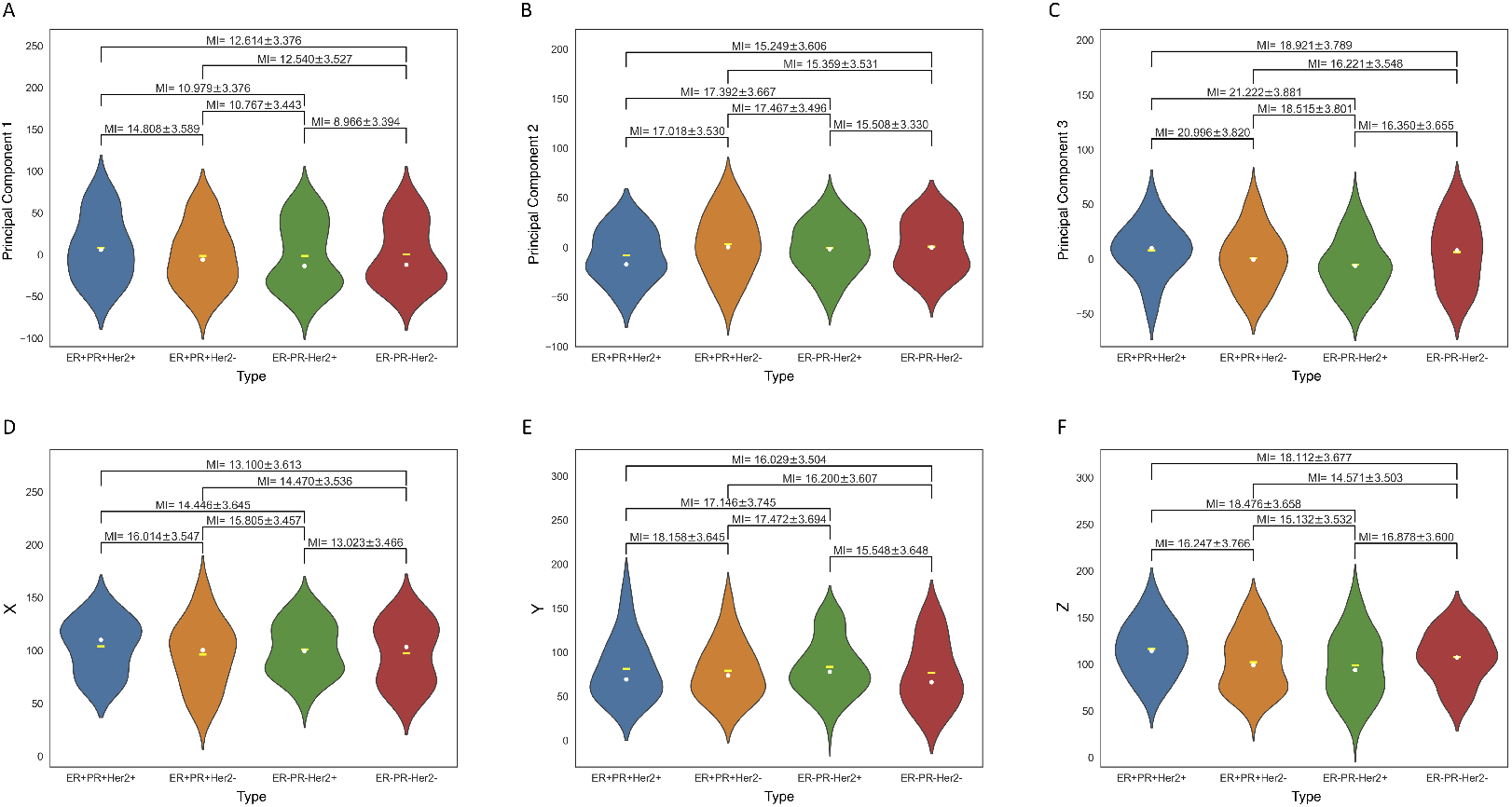
Violin plots (probability distribution functions) of metastatic distributions according to molecular groupings (indicated by color), comparing distributions in original Cartesian X-Y-Z coordinates, and Principal component coordinates (PC1-PC2-PC3). MI metric is shown for each pair. A) Distribution along PC1-axis according to molecular grouping. Yellow dash indicates mean, dot indicates median; C) Distribution along PC3-axis according to molecular grouping. Yellow dash indicates mean, white dot indicates median; D) Distribution along X-axis according to molecular grouping. Yellow dash indicates mean, white dot indicates median. We use this representation to arrange the subtypes from left to right in order of increasing divergence between the means and medians; E) Distribution along Y-axis according to molecular grouping. Yellow dash indicates mean, white dot indicates median; F) Distribution along Z-axis according to molecular grouping. Yellow dash indicates mean, white dot indicates median.

Figures 6 and 7 show the differences between anterior vs. posterior and lateral vs. medial lesions from the sagittal, axial, and coronal views (Figure 6) and according to molecular subtype groupings. While Figures 7 A-D show the Count (number of metastatic lesions), Figure 7 E-H shows the proportion in each of the regions. It is clear that from Figure 6, the majority of lesions are located in the posterior circulation or watershed areas, and BMBC are relatively rare in the anterior circulation. Figures 7 G,H demonstrate the differences in medial vs lateral distribution of these tumors. It is clear from Figure 7 G that midline tumors are most common across all molecular subtypes. In addition, it appears that Her2+ tumors have the highest proportion of medial metastases, and more rarely metastasize laterally. This is consistent (Figure 7 H) within the molecular subgroups as well, with ER+PR+Her2+ tumors having similar categorical distributions to ER-PR-Her2+ tumors but significantly different than TNBC or ER+PR+Her2-tumors.

**Fig. 6.**
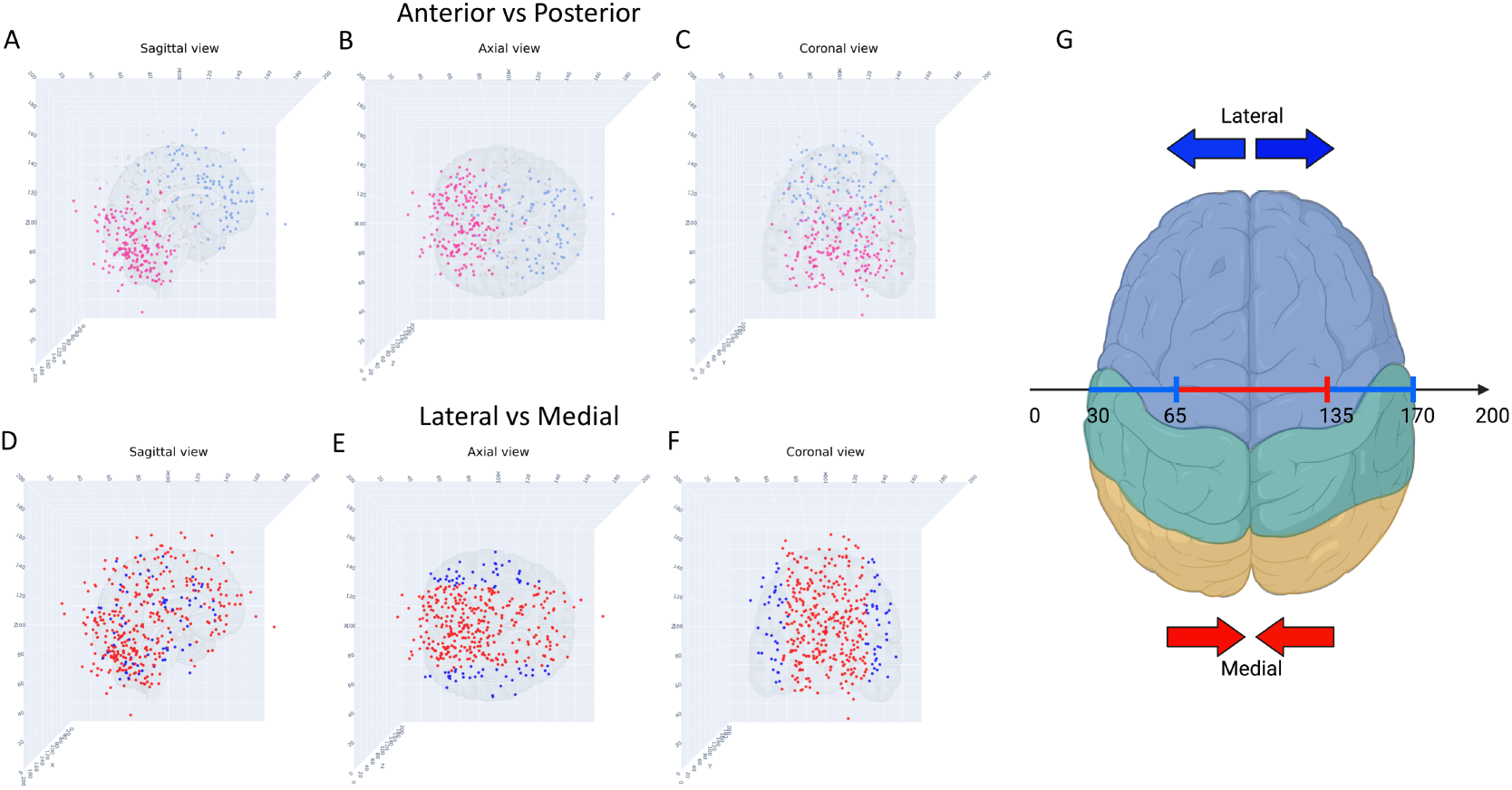
Scatter plots of metastatic tumors, anterior/posterior, lateral/medial, three different views. A) Sagittal view, blue indicates anterior, pink indicates posterior, grey dots indicate washout region; B) Axial view, blue indicates anterior, pink indicates posterior, grey dots indicate washout region; C) Coronal view, blue indicates anterior, pink indicates posterior, grey dots indicate washout region; D) Sagittal view, blue indicates lateral, red indicates medial; E) Axial view, blue indicates lateral, red indicates medial; F) Coronal view, blue indicates lateral, red indicates medial; G) Topographical illustration (axial) showing the X-coordinates corresponding to the lateral and medial regions.

**Fig. 7.**
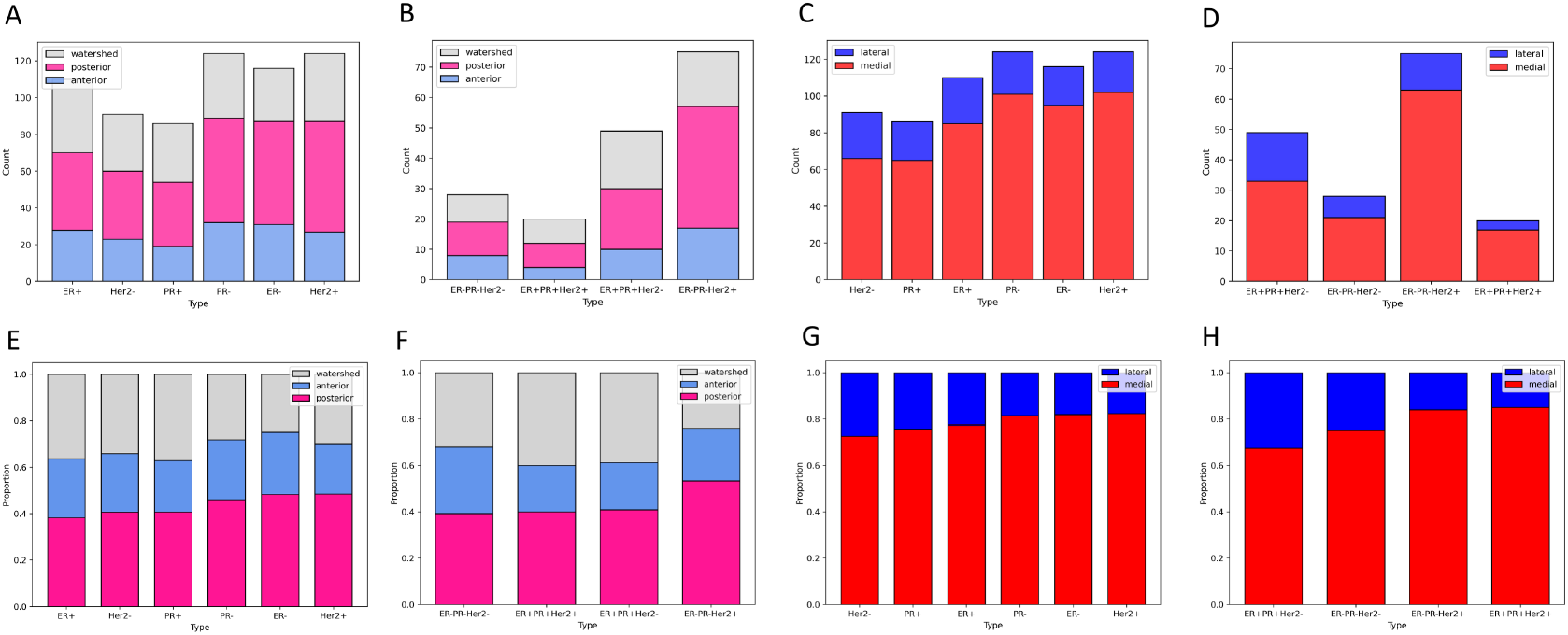
Histograms (Count and Proportion) showing spatial distribution (anterior/posterior; lateral/medial) by molecular subtypes and groupings. A) Count per molecular subtype, anterior (blue), posterior (pink), watershed (grey); B) Count per molecular grouping, anterior (blue), posterior (pink), watershed (grey); C) Count per molecular subtype, lateral (blue), medial (red); D) Count per molecular grouping, lateral (blue), medial (red); E) Proportion per molecular subtype, anterior (blue), posterior (pink), watershed (grey); F) Proportion per molecular grouping, anterior (blue), posterior (pink), watershed (grey); G) Proportion per molecular subtype, lateral (blue), medial (red); H) Proportion per molecular grouping, lateral (blue), medial (red).

**Fig. 8.**
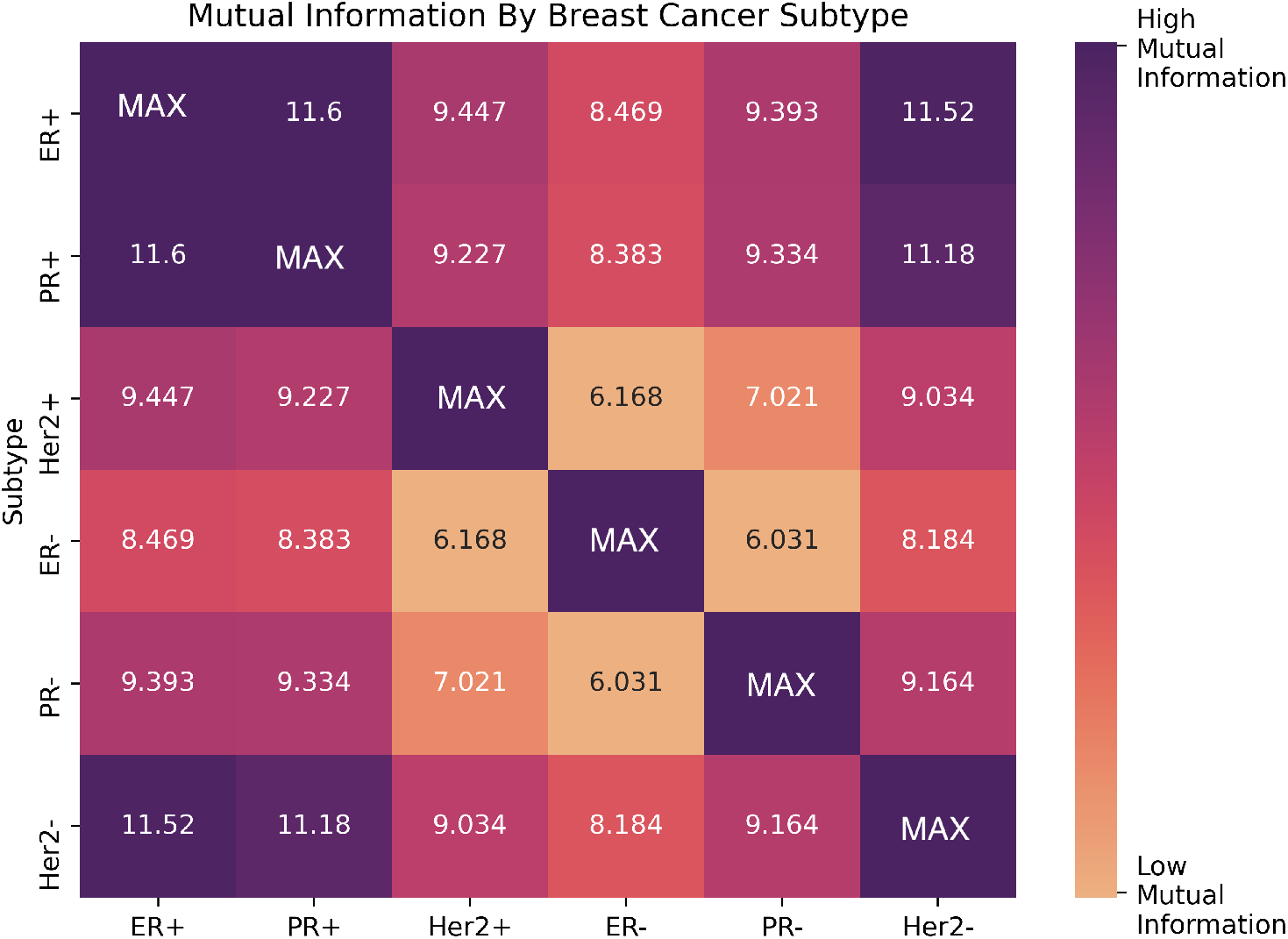
Mutual information heat map for the six breast cancer molecular subtypes along PC1-axis. Low mutual information indicates the distributions associated with the two subtypes are not highly dependent. High mutual information indicates the distributions associated with the two subtypes are highly dependent.

## 5. Discussion

Accurate quantitative characterization and analysis of BM distributions for primary breast cancer, broken down according to molecular subtypes, is an important step in the direction of highly personalized oncologic therapy and an understanding of the dynamics between BM subtypes and the TME that promote or inhibit the formation of metastasis. To further classify the relationship between a tumor and the microenvironment in which growth is facilitated or the genetic influences which allow for tumor growth in a particular environment, the specific location of tumor foci must be accurately and quantitatively analyzed. Although collecting, quantifying, and processing this information from large multicenter datasets is ongoing, our intention was to develop and share a practical and novel workflow for objective and data-driven analysis of BM distribution, along with useful quantitative techniques that are broadly applicable to other cancer types, larger data sets, and a wide range of centers whom intend to investigate similar relationships. While the seed-and-soil hypothesis has been an accepted overarching framework for over 100 years, detailed information about the spatial distributions of metastases in sensitive organs and broken down by tumor types and molecular subtypes is lacking [12]. In this study, we describe current methods for quantifying the spatial distribution of brain metastases, describe the utility of GKRS coordinates to facilitate this quantification, and discuss future applications and possibilities using widespread coordinate mapping and analysis.

In preliminary analyses, triple negative breast cancers or TNBC (i.e. estrogen receptor negative, progesterone receptor negative) with varying her2 status were the most spatially distinct. In contrast, hormone receptor positive tumors with differing her2 status were the most similar. This suggests that hormone receptor status may disproportionately influence the spatial distribution of metastases. One hypothesis is that hormone receptor status, when ‘silent’, then allows her2 status to drive spatial distribution of BM. Conversely, when ‘activated’ (e.g. progesterone positive and estrogen positive), differences in her2 status may be more muted, at least in the context of spatial distribution. Clinically, luminal breast cancer (hormone receptor positive, Her2 negative) demonstrates distinct responses to therapies, and have a slower rate of growth and more positive outcomes. In addition, there is a relationship between TNBC, Her2-negative/hormone receptor positive tumors and mutations in the genes BRCA1 and BRCA2. These additional genetic markers may influence the spatial makeup of these subtypes and may validate the mutual information scores determined between these subtypes. Furthermore, hormone receptor positive tumors, regardless of their Her2 status, tend to portend the best clinical outcomes for patients. While this phenomenon is currently largely driven by therapeutic targets afforded by hormone receptor positivity, there may be additional genetic drivers which also influence spatial distribution.

While several groups have aimed to categorize tumor location by subtype using MRIs, these studies are generally pilot studies and relatively small in sample size [19–22]. The non-granular level of anatomical precision from MRI studies (e.g. describing tumor location qualitatively as ‘frontal lobe’) often prevents further downstream analysis of these tumor distributions using advanced mathematical and computational means. This precision becomes important when discussing embryologic, signal-based and/or genetic and epigenetic influences in tumor development; discriminating between the midline frontal lobe and more lateral aspects is meaningful as these regions have different vascular distributions, functions and are likely embryologically driven by different mechanisms, despite being in the same lobe [22]. FOX genes, for instance, are theorized to drive midline brain development and Sonic Hedgehog (SHH) has been shown to drive cerebellar development [23–25]. The process of anatomical mapping of brain metastases when performed via MRI is also sensitive to variations in institutional MRI sequence protocol, and can influence the spatial mapping of tumors, as shown by a study by Kyeong et al [25] and Izustsu et al [26] who mapped genetic subtypes of breast cancer with differing MRI sequences and obtained conflicting results [25, 26]. Lastly, MRI reading requires a trained neuro-radiologist and is time consuming and tedious, preventing its widespread adoption. While advancements in machine learning and computer vision may allow for precise anatomical landmark distinction at scale, these techniques are not widespread [27].

Analysis using GKRS is a promising alternative to qualitative anatomical location analysis for a variety of reasons. GKRS Leksell coordinates are already collected at the time of radiosurgery and utilized in routine clinical care, allowing for ease of implementation. They are specific to each patient and each tumor and provide accurate, three-dimensional coordinates of tumor centroids. Finally, GKRS data are easily scalable and standardizable across institutions for future data collection and does not require manual annotation by skilled professionals, and can be analyzed in an objective and quantitative fashion rather than using categorical descriptors, thereby increasing internal validity of the analyses.

By transforming the data from the original Leksell anatomical coordinates to the principal coordinate axes, we are using an optimal data-derived coordinate system that highlights the axis along which there is the largest spread (PC1), the second largest spread (PC2), and the least spread (PC3) of the data. What we lose in this linear transformation is an easily interpretable anatomical frame, but we gain the ability to quantify what would otherwise be very subtle differences among molecular subgroups. We have retained the original anatomical frame, however, to depict the kernel density plots showing the clustering regions along the 3 two-dimensional projections, in order to more easily discern the physical locations in the brain where the clusters occur and to correlate this with blood flow patterns.

We further demonstrate that the results obtained by the GKRS coordinate spatial distribution system are accurate and can elucidate meaningful differences in molecular subtype distribution patterns. It has been well described that breast cancer preferentially metastasizes to the cerebellum; KDE plots from GKRS data demonstrate the preference for the posterior circulation and below the central cranio-caudal axis, consistent with a cerebellar distribution [1, 21]. Izutsu et al [26]found that in their cohort of 67 patients with 437 tumors, Her2 positivity was associated with metastases in the putamen and thalamus and less frequently in the cerebellum [26]. Figure 7 corroborates these findings, wherein Her2+ tumors appear to be preferentially distributed on the midline (thalamus and putamen are midline structures). Kyeong et al [25] found that TNBC was evenly distributed in the brain, which is supported by Figure 7 F, where TNBC appears to have a relatively uniform distribution between anterior, posterior and watershed areas of circulation [25]. It is important to note that our study did not corroborate all of the findings within the literature-for example Kyeong et al [25] contradicted the findings by Izutsu et al [26] (and our analysis) and found BM from Her2 positive and luminal type tumors more common in the cerebellum and occipital lobe. These inconsistencies (and differences in sequence methods) highlight the need for high quality, standardized data collection and analysis methods. Using mutual information, data on subtype similarity may be explored: for instance, TPBC and hormone negative BC (TNBC, ER-PR-Her2+) had two of the most divergent patterns of distribution. This supports known characterization of BC, where hormone receptor positivity portends significantly improved outcomes [28]. Further characterization of and groupings of subtypes with higher MI coefficients (higher similarity) should also be explored (with larger data sets), such as between ER+PR+Her2+ tumors and ER+PR+Her2-tumors; it may be that the clustering of these tumors are both relatively non-preferential, hence they have high MI coefficients, however there may be underlying factors related to tumor microenvironment or other genes which may drive tumorigenesis in similar locations. Subsequent translational/animal models which attempt to categorize growth of tumors based on their location should prioritize investigating tumor subtypes with the most convergent and divergent MI indices.

### 5.1 Opportunities for Advancement in Diagnosis and Treatment

Neurotransmitters (e.g. gamma-aminobutyric acid (GABA), glutamate, dopamine, etc) are the biochemical backbone for synaptic signaling, but are also utilized for other cellular functions. These neurotransmitters are present in varying concentrations in different regions; for example, GABA-ergic communication is predominant in cerebellum. This difference is also highlighted by blood-flow; and it is speculated that BM have a predominance in the cerebellum due to the difference in blood flow to those regions, however it is unknown why this affect has a nonuniform impact across primary cancers and subtypes. Understanding the spatial distribution of BM based on molecular subtype may further characterize tumor ability to adapt to regional microenvironments based on these neurotransmitter distributions, and may promote BM progression [3, 10,29]. There is a need for large, multi-center studies which utilize standardized data collection criteria to accurately map our brain metastases to avoid inaccuracies as previously mentioned, and enhance generalizability and external validity of this work. In addition, the current advantage of MRI mapping vs GKRS is the ability to develop a 1-1 anatomic map. Accordingly, efforts should be made to create a Leksell-Anatomic mapping, wherein specific X,Y,Z coordinates map to a specific location on a standardized cartesian plane. These mapping classifications must be corroborated with in-vitro and animal models, demonstrating the ability to seed tumor more readily in certain areas of the brain, or identify DNA/RNA lineages specific to tumor locations. Finally, this data must be correlated with clinical factors (e.g. time to diagnosis, overall survival, etc.) which can allow for the development of clinical decision trees. Groups have postulated that the accurate classification of subtypes and correlation with high-risk subgroups might warrant increased surveillance in the period following cancer diagnosis but before BM diagnosis, or even prophylactic, low dose radiation to regions of the brain with high susceptibility [26]. These clinical implementations remain distant, however the systematic, quantifiable mapping of BM distributions is an important first step in personalized oncologic care for the patient with BMBC.

### 5.2 Limitations

There are limitations to the current study. While stereotactic headsets are standardized in their size, they are fit to a patient’s specific head size which may introduce variation in coordinate recordings. Studied across a cohort of hundreds or thousands of patients, however, these individual cranial-frame variations are likely to normalize and not preclude meaningful statistical comparison. Secondly, the anatomical distributions demonstrated (anterior/posterior, medial/lateral) are Cartesian-derived and may have a limited degree of inaccuracy, although GKRS accuracy has been reported to be on the order of 1mm. The data itself introduces a level of systematic bias as it only accounts for patients who had GKRS for treatment of BM, and not patients who elect not to undergo GKRS, those who undergo whole brain radiation, or have undiagnosed BMBC. Furthermore, correlation with MRI endpoints would significantly strengthen this work. However, advanced imaging studies which may allow us to make more definitive claims regarding the tumor-tumor microenvironment specific to anatomic endpoints (e.g. MR angiograms, perfusion MRI, tractography, etc.) were not performed systematically across any significant subset of patients. Lastly, given that individual cancers themselves have differential distribution patterns, by definition, variance within cancers will be far more subtle. Accordingly, our samples may be significantly underpowered to detect meaningful difference in cancer subtype distribution, which is why we employ the bootstrap/re-sampling method. Scaling the analysis described using the current workflow to thousands of BMK patients from multi-center consortia will increase power and allow more meaningful and granular comparison of cancer and molecular BM subtypes.

## 6. Conclusion

We demonstrate a novel, objective, data-based methodology for classifying and analyzing the spatial distribution of brain metastases by breast cancer molecular subtypes using stereotactic coordinates, principal component coordinates (PC), and kernel density estimators (KDE) to highlight clustering regions in the brain. We then compare distributions associated with different molecular subtypes using the mutual information (MI) metric, which is a widely used bioinformatic metric [15, 16], but to our knowledge has not been used in the current context. This systematic, quantitative method for classifying BM distribution is easy to scale, accurate, and a meaningful step forward towards understanding the relationship between BM tumor microenvironment and tumorigenesis. Her2+ vs. Her2-cancers may show differential patterns based on this pilot study data and novel methodology.

## Data Availability

All data produced in the present work are contained in the manuscript.

## Acknowledgments

Partial funding through the USC Norris Comprehensive Cancer Center’s Multi-Level Cancer Risk Prediction Models pilot project award, ‘Molecular, Clinical and Neuro-imaging Determinants of Spatiotemporal Pathogenesis of Cancer-Specific Brain Metastases: Data Analysis and Longitudinal Modeling’ (12/01/2020-11/30/2021) is gratefully acknowledged.

---

**Fig. S9.**
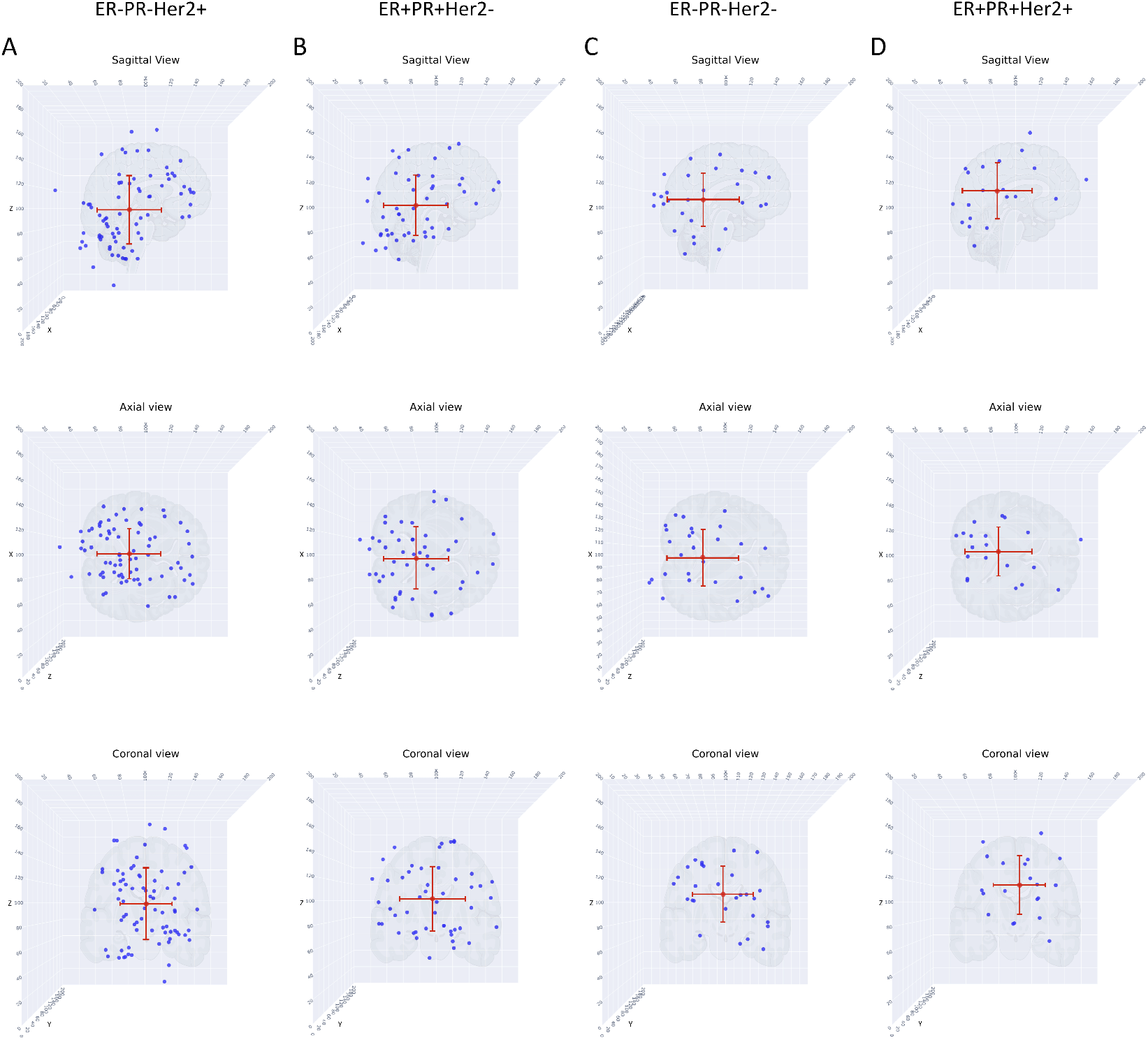
Scatter plots of metastatic tumor distributions according to genetic subgroups, sagittal, axial, and coronal views. Red dot indicates mean. A) Column showing ER-/PR-/Her2+ subgroup, three views; B) Column showing ER+/PR+/Her2-subgroup, three views; C) Column showing TNBC subgroup, three views; D) Column showing TPBC subgroup, three views.

**Fig. S10.**
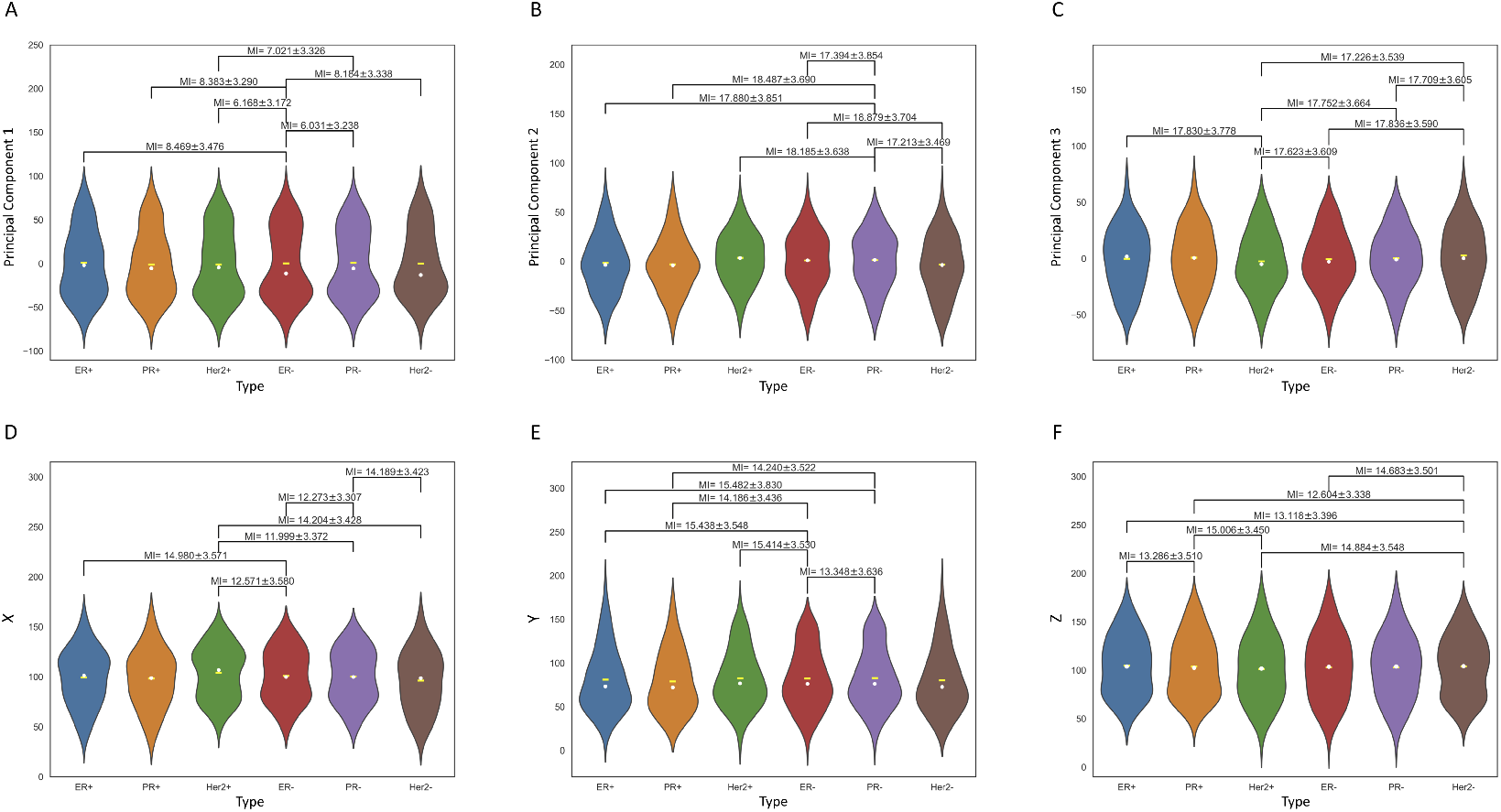
Violin plots (probability distribution functions) of metastatic distributions according to molecular subtype (indicated by color), comparing distributions in original Cartesian X-Y-Z coordinates, and Principal component coordinates (PC1-PC2-PC3). MI metric is shown for each pair. A) Distribution along PC1-axis according to molecular subtype. Yellow dash indicates mean, white dot indicates median; B) Distribution along PC2-axis according to subtype. Yellow dash indicates mean, white dot indicates median; C) Distribution along PC3-axis according to subtype. Yellow dash indicates mean, white dot indicates median; D) Distribution along X-axis according to molecular subtype. Yellow dash indicates mean, white dot indicates median. We use this representation to arrange the subtypes from left to right in order of increasing divergence between the means and medians; E) Distribution along Y-axis according to molecular subtype. Yellow dash indicates mean, white dot indicates median; F) Distribution along Z-axis according to molecular subtype. Yellow dash indicates mean, white dot indicates median.

**Table S1.**
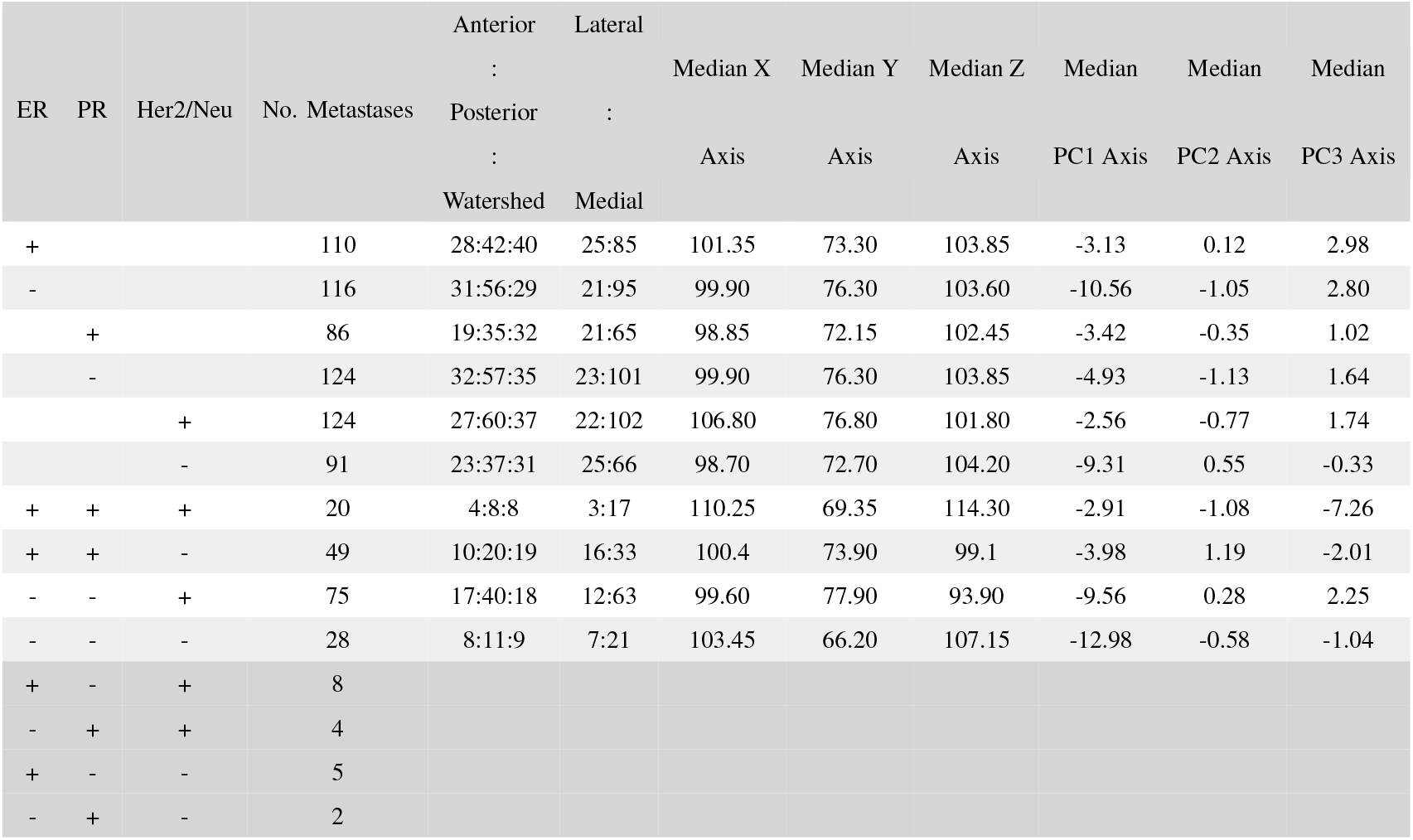
Number of brain metastases and proportion of different spatial subgroupings along with the medians in Cartesian and Principal Component coordinates by tumor subtype. The last four molecular subgroupings (last four rows) are not considered in this paper due to the small number of data points.

**Table S2.** Mutual Information. Ranked listing (from smallest to largest) of MI between pairs of molecular subtypes along each of the coordinate axes, using the PC1 coordinate axis values to order the list. Smaller MI values indicate weaker mutual dependence (i.e. more independence), larger MI values indicate stronger mutual dependence.

|  | PC1 | PC2 | PC3 | X | Y | Z |
| --- | --- | --- | --- | --- | --- | --- |
| ER-PR-Her2+/<br>ER-PR-Her2- | 8.966 ± 3.394 | 15.508 ± 3.330 | 16.350 ± 3.655 | 13.023 ± 3.466 | 15.548 ± 3.648 | 16.878 ± 3.600 |
| ER+PR+Her2-/<br>ER-PR-Her2+ | 10.767 ± 3.443 | 17.467 ± 3.496 | 18.515 ± 3.801 | 15.805 ± 3.457 | 17.472 ± 3.694 | 15.132 ± 3.532 |
| ER+PR+Her2+ /<br>ER-PR-Her2+ | 10.979 ± 3.376 | 17.392 ± 3.667 | 21.222 ± 3.881 | 14.446 ± 3.645 | 17.146 ± 3.745 | 18.476 ± 3.658 |
| ER+PR+Her2-/<br>ER-PR-Her2- | 12.540 ± 3.527 | 15.359 ± 3.531 | 16.221 ± 3.548 | 14.470 ± 3.536 | 16.200 ± 3.607 | 14.571 ± 3.503 |
| ER+PR+Her2+/<br>ER-PR-Her2- | 12.614 ± 3.376 | 15.249 ± 3.606 | 18.921 ± 3.789 | 13.100 ± 3.613 | 16.029 ± 3.504 | 18.112 ± 3.677 |
| ER+PR+Her2+ /<br>ER+PR+Her2- | 14.808 ± 3.589 | 17.018 ± 3.530 | 20.996 ± 3.820 | 16.014 ± 3.547 | 18.158 ± 3.645 | 16.247 ± 3.766 |
| PR- / ER- | 6.031 ± 3.238 | 17.394 ± 3.854 | 17.993 ± 3.653 | 12.273 ± 3.307 | 13.348 ± 3.636 | 17.442 ± 3.644 |
| Her2+ / ER- | 6.168 ± 3.172 | 19.844 ± 3.765 | 17.623 ± 3.609 | 12.571 ± 3.580 | 15.414 ± 3.530 | 17.102 ± 3.598 |
| Her2+ / PR- | 7.021 ± 3.326 | 18.185 ± 3.638 | 17.752 ± 3.664 | 11.999 ± 3.372 | 15.715 ± 3.678 | 17.683 ± 3.720 |
| Her2- / ER- | 8.184 ± 3.338 | 18.879 ± 3.704 | 17.836 ± 3.590 | 14.506 ± 3.569 | 16.038 ± 3.762 | 14.683 ± 3.501 |
| PR+ / ER- | 8.383 ± 3.290 | 20.139 ± 3.885 | 18.164 ± 3.536 | 15.609 ± 3.565 | 14.186 ± 3.436 | 15.196 ± 3.607 |
| ER+ / ER- | 8.469 ± 3.476 | 19.238 ± 3.652 | 17.926 ± 3.622 | 15.484 ± 3.515 | 15.438 ± 3.548 | 15.482 ± 3.679 |
| Her2+ / Her2- | 9.034 ± 3.363 | 19.914 ± 3.792 | 17.226 ± 3.539 | 14.204 ± 3.428 | 18.322 ± 3.758 | 14.884 ± 3.548 |
| PR- / Her2- | 9.164 ± 3.319 | 17.213 ± 3.469 | 17.709 ± 3.605 | 14.189 ± 3.423 | 15.927 ± 3.691 | 15.359 ± 3.520 |
| Her2+ / PR+ | 9.227 ± 3.452 | 21.094 ± 3.730 | 18.118 ± 3.514 | 15.350 ± 3.506 | 16.584 ± 3.661 | 15.006 ± 3.450 |
| PR+ / PR- | 9.334 ± 3.323 | 18.487 ± 3.690 | 18.253 ± 3.550 | 15.325 ± 3.542 | 14.24 ± 3.522 | 15.662 ± 3.541 |
| ER+ / PR- | 9.393 ± 3.582 | 17.88 ± 3.851 | 18.09 ± 3.490 | 14.980 ± 3.571 | 15.482 ± 3.830 | 16.098 ± 3.715 |
| Her2+ / ER+ | 9.447 ± 3.351 | 20.288 ± 3.861 | 17.830 ± 3.778 | 15.093 ± 3.558 | 17.836 ± 3.753 | 15.588 ± 3.586 |
| PR+ / Her2- | 11.178 ± 3.466 | 20.357 ± 3.804 | 17.978 ± 3.604 | 17.838 ± 3.564 | 16.798 ± 3.715 | 12.604 ± 3.338 |
| ER+ / Her2- | 11.516 ± 3.469 | 19.579 ± 3.714 | 17.827 ± 3.669 | 17.216 ± 3.831 | 18.183 ± 3.745 | 13.118 ± 3.396 |
| ER+ / PR+ | 11.601 ± 3.455 | 20.712 ± 3.723 | 18.224 ± 3.532 | 18.455 ± 3.691 | 16.696 ± 3.625 | 13.286 ± 3.510 |

